# Frontier Large Language Models for Comprehensive Medication Review in CKD Patients with Polypharmacy: A Trap-Embedded Synthetic Benchmark

**DOI:** 10.64898/2026.05.23.26353939

**Authors:** Kai-Chou Chuang, Hsuan-Jen Lin, Hsuan-Ming Lin

## Abstract

**Background:** Patients with CKD and polypharmacy face high rates of drug-related problems, yet comprehensive medication review remains time-intensive and inconsistently performed. Large language models (LLMs) may augment this process, but existing benchmarks use multiple-choice formats that do not reflect open-ended, nephrology-specific review. We developed a trap-embedded synthetic CKD benchmark and evaluated five current-generation LLMs (GPT-5.4, Claude Sonnet 4.6, Gemini 3.1 Pro, Grok 4.1 Fast, DeepSeek R1; tested April–May 2026) for open-ended medication review.

**Methods:** Fifty synthetic CKD cases across three complexity groups (G3a–G3b [*n* = 20], G4 [*n* = 15], G5/G5D/transplant [*n* = 15]) with 8–12 medications and ≥2 embedded clinical traps each were scored against nephrologist-adjudicated gold standards. Each model produced three independent responses per case (temperature 0; 750 total outputs). Primary endpoint was per-case macro F1; secondary endpoints were safety-critical omission rate, PI-adjudicated hallucination rate, and intra-model consistency. Blinded inter-rater reliability for gold-standard item detection was assessed on a 30% sample.

**Results:** Consensus-level macro F1 ranged from 0.41 (Claude Sonnet 4.6) to 0.49 (Grok 4.1 Fast) (Friedman *P* < 0.001). Phosphate binder timing (11%) and hyperkalemia combinations (33%) were poorly detected across all models. Safety-critical omission rate ranged from 22% to 48% (*P* < 0.001); PI-adjudicated hallucination ranged from 0% (GPT-5.4) to 54% (DeepSeek R1), including fabricated dose caps and non-existent guideline citations. Blinded reliability for gold-standard item detection was high (*κ* = 0.934, *n* = 92).

**Conclusions:** This nephrology-specific benchmark exposes clinically important LLM blind spots that generic multiple-choice evaluations would not detect. Heterogeneous hallucination and omission rates indicate that model selection and domain-specific guardrails should precede any clinical deployment of LLM-assisted CKD medication review. Prospective validation with real patient data and human comparators is required before deployment recommendations can be made.

**Key Points:**

- All five LLMs poorly detected phosphate binder timing (11%) and hyperkalemia combinations (33%) in nephrology-specific medication review.
- Macro F1 ranged 0.41 to 0.49, safety-critical omissions 22–48%, and hallucination 0–54%, with qualitatively distinct error patterns.
- Open-ended scoring exposed LLM failures invisible to multiple-choice benchmarks, supporting specialty evaluation before deployment.

## 1 Introduction

Current clinical decision support tools detect contraindicated drug combinations through rule-based lookup but cannot determine whether a patient’s calcium carbonate is taken with meals— the distinction between effective phosphate binding and mere calcium supplementation in CKD. This class of administration-context judgment, and analogous multi-drug integration problems requiring specialist synthesis, defines the clinical substance of comprehensive medication review (CMR)—the systematic evaluation of an entire regimen for appropriateness, safety, and completeness [1, 2]. Over 800 million people are affected by CKD globally [3, 4], and those in stages G3–G5 typically manage 8–12 concurrent medications [5, 6]—a high-stakes environment in which drugs require eGFR-adjusted dosing [7], nephrotoxic agents must be identified, and guideline-recommended therapies initiated at stage-appropriate thresholds [8, 9]. Pharmacist-led CMR in non-CKD populations identifies 2–5 clinically significant drug-related problems per patient yet misses 20–40% of problems detectable by specialist review [10–12]—a performance gap that may be wider in nephrology, where subspecialty knowledge is required.

Large language models (LLMs) have demonstrated strong performance on medical knowledge assessments [13, 14] and show potential for medication-related tasks including interaction checking and dose adjustment [15–17]. However, three critical gaps limit the translational relevance of existing evidence for nephrology practice.

First, the *model generation gap*: most published evaluations tested GPT-4 or Claude 3 (2023–2024 generation). The 2025–2026 frontier — GPT-5.4, Claude Sonnet 4.6, Gemini 3.1 Pro, Grok 4.1 Fast, and open-source models such as DeepSeek R1 — remains unevaluated for clinical medication review [18].

Second, the *specialty-specific context gap*: CKD medication management involves nuances rarely captured in general-medicine benchmarks, including stage-dependent dose thresholds, dialyzability, transplant immunosuppressant interactions, and administration timing constraints (e.g., phosphate binders relative to meals). ChatGPT achieved only 16.7% accuracy for renal dose interventions in one evaluation [19], yet prior LLM polypharmacy studies have focused predominantly on geriatric or general-medicine populations [20].

Third, and most critically, the *task format gap*: nearly all existing evaluations employ multiple-choice questions or closed-ended prompts, where LLM performance drops substantially when converted to open-ended formats [21]. Real-world CMR is fundamentally an open-ended generative task in which the clinician must independently identify all drug-related problems without predefined answer choices. This format mismatch inflates apparent performance and limits clinical actionability.

To address these gaps, a nephrology-specific benchmark was developed comprising 50 synthetic CKD cases with 10 categories of embedded clinical traps, nephrologist-validated gold standards, and 13 prespecified scoring decision rules. Five contemporary LLMs—representing the four leading commercial providers and the top-performing open-source reasoning model as of April 2026—were each evaluated on open-ended CMR tasks mirroring clinical workflow, with independent blinded inter-rater reliability assessment (*κ* = 0.934). This study characterizes the detection capability and safety profile of current-generation LLMs for CKD-specific medication review.

## 2 Methods

This cross-sectional study is reported following STROBE guidelines [22] (Supplementary Appendix D), with additional AI-specific reporting elements adapted from CONSORT-AI [23] where applicable.

### 2.1 Study Design

We conducted a cross-sectional evaluation of LLM-generated comprehensive medication reviews against nephrologist-adjudicated gold standards using 50 synthetic CKD cases. The study was approved by the An-Nan Hospital Research Ethics Committee under umbrella protocol 113TMANH-REC011(CR-1), which covers this research program. The present Phase A analysis used only fully synthetic cases constructed from public guidelines and contained no identifiable patient data; the same umbrella approval also covers the planned Phase B chart-review extension with real patient data.

### 2.2 Synthetic Case Construction

Fifty cases were distributed across three complexity groups: Group A (basic; CKD G3a–G3b, *n* = 20, 8 medications), Group B (advanced; G4, *n* = 15, 10 medications), and Group C (complex; G5/G5D/transplant, *n* = 15, 12 medications). Cases were constructed by a nephrologist (H-M.L.) referencing KDIGO 2024 guidelines [8], STOPP/START version 3 [24], and publicly available renal dosing references. Each case contained ≥2 embedded clinical traps drawn from 10 prespecified categories (T01–T10; Supplementary Table S1), covering contraindications, dose adjustment errors, drug–drug interactions, deprescribing opportunities, missing therapies, and administration timing errors. Nine trap categories appeared in ≥8 cases per protocol; T09 (transplant drug interactions) was limited to the 5 transplant cases by clinical design. During post-construction validation, 13 cases (26%) had *>*30% discrepancy between stated eGFR and serum creatinine via CKD-EPI 2021 reverse calculation; SCr values were corrected while preserving the stated eGFR and CKD stage. Sensitivity analysis confirmed stable results (ΔF1 = 0.010; Supplementary Table S7).

### 2.3 Gold Standard Development

For each case, a comprehensive list of expected findings was prespecified by the PI (H-M.L., nephrologist), referencing KDIGO guidelines, UpToDate drug dosing databases, and Lexicomp interaction databases (2026 Q1 versions). Each gold-standard problem was classified by type and assigned a severity level (0–4). Thirteen operational decision rules (DR-01 through DR-13) and four hallucination rules (HR-01 through HR-04) were finalized before scoring began (Supplementary Appendix B; April 25, 2026). Gold-standard completeness was reviewed by an independent nephrologist (H-J.L.) on a stratified 30% sample (15/50 cases). The completeness reviewer was provided with only the case description and the existing gold-standard list; the reviewer was *blinded* to all LLM responses, model identities, and aggregate scoring results during this review. The review identified 22 additional problems, 12 items requiring revision, and 1 item for removal or downgrade (37 actions across 12 cases). Added problems were distributed across severity levels (predominantly Level 2–3 moderate severity; none Level 4 critical) and across all case groups, rather than concentrated in any single model’s strength or weakness. After the update, the complete set of LLM responses was re-scored against the updated gold standard (commit-tagged “gold-standard completeness update + full rerun,” May 10, 2026); all reported results use the post-update gold standard. The final updated gold standard contained 307 scored problems across 50 cases.

### 2.4 Large Language Models

Five contemporary LLMs were selected to represent four major commercial providers and a leading open-source reasoning model as of April 2026: GPT-5.4 (OpenAI), Claude Sonnet 4.6 (Anthropic), Gemini 3.1 Pro (Google), Grok 4.1 Fast (xAI), and DeepSeek R1 (DeepSeek; open-source, accessed via OpenRouter). All models were tested at temperature 0 with a standardized system prompt (Supplementary Appendix A). Three independent API calls were made per case per model (750 total outputs). Exact API model identifiers, access routes, testing dates, and per-model token budgets are detailed in Supplementary Table S9.

Model configurations differed in four respects that may influence between-model comparison and should be considered when interpreting results. First, GPT-5.4, Claude Sonnet 4.6, and Gemini 3.1 Pro were tested in their standard production modes; Grok 4.1 Fast was tested in non-reasoning mode (the xAI tier without reasoning trace); DeepSeek R1 is an inherently reasoning model whose native reasoning trace was included within the output token budget (the reasoning trace was excluded from scoring; only the final answer content was parsed). Second, GPT-5.4 and Gemini 3.1 Pro were accessed via provider-native APIs; Claude Sonnet 4.6, Grok 4.1 Fast, and DeepSeek R1 were accessed via OpenRouter, with the exact DeepSeek R1 checkpoint determined by OpenRouter routing at query time. Third, testing windows differed: GPT-5.4, Gemini 3.1 Pro, and Grok 4.1 Fast were tested April 26–28, 2026; DeepSeek R1 was tested May 3, 2026; Claude Sonnet 4.6 was re-tested on May 17, 2026 after a methodological update (described below). Fourth, maximum output tokens differed: GPT-5.4, Gemini 3.1 Pro, Grok 4.1 Fast, and DeepSeek R1 used 8,192 tokens (no output reached this ceiling); Claude Sonnet 4.6 used 16,384 tokens after the May 17 re-test (rationale below). These configuration asymmetries are documented in Supplementary Table S9 and revisited in the Discussion as a factor limiting strict inter-model ranking.

#### Provenance note: Claude Sonnet 4.6 retest at 16,384 tokens (May 17, 2026)

An initial Claude Sonnet 4.6 test on April 26–28 at 8,192 maximum output tokens exhibited a 29% rate of output truncation at the token limit (60% in Group C complex cases), with responses cut off mid-table or mid-sentence. Because this truncation pattern was systematically more frequent in complex cases, it could bias Claude Sonnet 4.6’s apparent detection metrics. We therefore re-tested Claude Sonnet 4.6 on May 17, 2026 at 16,384 maximum output tokens via OpenRouter (the same underlying model as the original direct-API run). All 150 retest outputs completed without truncation; maximum observed output was 12,981 tokens, with mean 7,877 tokens. All Claude Sonnet 4.6 results reported here use the May 17 retest data; outputs from the other four models retained their April–May 2026 original runs. The scoring pipeline (extraction, gold-standard matching, hallucination sweep, DR-06 reclassification) was re-run on the new Claude Sonnet 4.6 outputs.

### 2.5 Scoring Pipeline

A two-stage approach balanced efficiency with accuracy. In Stage 1, a dedicated extraction model (Claude Sonnet 4.5; temperature 0; full extraction prompt in Supplementary Appendix A) parsed each free-text response into structured problem tuples (drug, issue_type, action), with candidate matching against the gold standard using decision rules DR-01–DR-13. Extraction accuracy was validated on a 60% stratified random sample (333 tuples), achieving 100% agreement with PI manual verification. An independent second nephrologist confirmed 98% extraction completeness on 50 raw outputs (10 per model). To assess potential bias from the shared architecture between the extraction model (Claude Sonnet 4.5) and one tested model (Claude Sonnet 4.6), extraction accuracy was examined separately for Claude-generated responses; no difference was observed (Supplementary Table S6).

In Stage 2, the PI reviewed all candidate classifications, confirming or overriding TP/FP/FN designations, with 14 corrections applied. A hallucination sweep was performed for each response following rules HR-01–HR-04. The Stage 2 PI adjudication was performed with model identity *visible* to the PI; this is acknowledged as a residual bias-risk limitation (Discussion) and was mitigated by the prespecified decision rules and the independent blinded inter-rater reliability assessment described below.

### 2.6 Sensitivity Analysis (DR-06)

Decision Rule 06 addressed clinically valid recommendations identified by LLMs that fall outside the gold-standard scope. The PI classified all false positives into true errors, clinically valid extras (DR-06), or theoretically correct but clinically insignificant (DR-07), based on aggregated (drug, issue_type) patterns to provide partial blinding to model identity. Extended F1 was computed excluding DR-06 and DR-07 items as an exploratory upper-bound estimate.

### 2.7 Intra-Model Consistency and Inter-Rater Reliability

For each case, pairwise Jaccard similarity was computed on (drug, issue_type) tuples across the three runs. Consensus scoring used majority rule (≥2/3); when all three runs differed, Run 1 was used as primary.

A second nephrologist (H-J.L.), blinded to the primary scorer’s decisions, independently scored a stratified 30% sample (15 cases) using the same decision rules. Cohen’s *κ* was calculated at the problem level (TP vs FN classification per gold-standard item).

### 2.8 Statistical Analysis

All primary analyses were conducted at the model-case consensus level (50 consensus evaluations per model; 250 total). The primary endpoint was macro F1 (per-case harmonic mean of precision and recall, averaged across cases). Bootstrap 95% confidence intervals (*n* = 2,000 resamples) were used for macro metrics; Wilson score intervals for binary proportions (SCE rate, hallucination rate). The Friedman test compared F1 across five models; Cochran’s Q compared SCE rates. Pairwise post-hoc tests used Wilcoxon signed-rank (F1) and McNemar (SCE) with Bonferroni correction (*α*_adj_ = 0.005 for 10 pairs). All analyses used Python 3.11+ (scipy, statsmodels, pandas).

## 3 Results

### 3.1 Case Characteristics

Table 1 summarizes the 50 synthetic cases. Mean age was 67.5 ± 9.1 years (68% male), with mean eGFR 29.7 ± 15.4 mL/min/1.73m^2^ and 9.8 ± 1.7 medications per case. Group C included 5 dialysis and 5 transplant cases. A total of 307 gold-standard problems were distributed across cases after independent completeness review (mean 6.1 per case; range 2–13).

**Table 1.** Characteristics of 50 synthetic CKD cases by complexity group.

|  | Group A<br>(*n* = 20) | Group B<br>(*n* = 15) | Group C<br>(*n* = 15) | All<br>(*N* = 50) |
| --- | --- | --- | --- | --- |
| CKD Stage | G3a–G3b | G4 | G5/G5D/Tx | — |
| Age, years | 67.2 ± 9.6 | 63.2 ± 8.1 | 72.1 ± 7.6 | 67.5 ± 9.1 |
| Male sex, % | 70.0 | 66.7 | 66.7 | 68.0 |
| eGFR, mL/min/1.73m<sup>2</sup> | 44.0 ± 9.2 | 22.6 ± 3.5 | 17.8 ± 14.2 | 29.7 ± 15.4 |
| Medications, mean | 8.0 | 10.0 | 12.0 | 9.8 ± 1.7 |
| Comorbidities, mean | 2.7 ± 0.7 | 4.3 ± 0.9 | 5.1 ± 0.7 | 3.9 ± 1.3 |
| Dialysis, % | 0 | 0 | 33.3 | 10.0 |
| Transplant, % | 0 | 0 | 33.3 | 10.0 |

Values: mean ± SD or %. Tx = transplant. eGFR estimated by CKD-EPI 2021. Group C large SD reflects intentional heterogeneity (G5 non-dialysis, G5D, and post-transplant with variable graft function).

### 3.2 Overall Model Performance

Consensus-level macro F1 ranged from 0.41 (Claude Sonnet 4.6; 95% CI: 0.37–0.45) to 0.49 (Grok 4.1 Fast; 95% CI: 0.45–0.53), with significant heterogeneity across models (Friedman *χ*^2^ = 20.46, *df* = 4, *P* = 0.0004; Table 2). Recall was highest for Claude Sonnet 4.6 (0.75) and GPT-5.4 (0.73), corresponding to longer mean response lengths that flagged a larger number of candidate problems. Precision was highest for Grok 4.1 Fast (0.42) and Gemini 3.1 Pro (0.39), reflecting more selective reporting. Significant pairwise differences (Bonferroni *α* = 0.005) were observed for Claude Sonnet 4.6 vs Gemini 3.1 Pro (*P* = 0.0006), Claude Sonnet 4.6 vs Grok 4.1 Fast (*P* = 0.0005), and Grok 4.1 Fast vs DeepSeek R1 (*P* = 0.0010).

**Table 2.** Primary outcomes per model across all 50 cases.

| Model | Recall | Precision | F1 | SCE | Hall. |
| --- | --- | --- | --- | --- | --- |
| GPT-5.4 | .73 (.67–.78) | .32 (.29–.36) | .44 (.39–.48) | .22 (.13–.35) | .00 (.00–.07) |
| Claude Sonnet 4.6 | .75 (.69–.80) | .29 (.26–.33) | .41 (.37–.45) | .22 (.13–.35) | .18 (.10–.31) |
| Gemini 3.1 Pro | .63 (.57–.69) | .39 (.36–.43) | .47 (.43–.51) | .40 (.28–.54) | .12 (.06–.24) |
| Grok 4.1 Fast | .62 (.56–.69) | .42 (.38–.45) | .49 (.45–.53) | .28 (.17–.42) | .52 (.39–.65) |
| DeepSeek R1 | .52 (.46–.59) | .37 (.32–.42) | .42 (.37–.47) | .48 (.35–.61) | .54 (.40–.67) |
| Friedman $\chi^2$ (F1): 20.46, $P = 0.0004$ | | | Cochran’s Q (SCE): 23.10, $P = 0.0001$ | | |

**Primary endpoint: macro F1** (per-case harmonic mean of precision and recall, averaged across 50 cases). Values: point estimate (95% CI). CI: bootstrap ( $n = 2,000$ ) for macro recall, precision, F1; Wilson for SCE, hallucination. SCE = safety-critical error (cases with $\geq 1$ Level 3–4 missed problem, i.e., omission-type error; commission-type errors are reported separately as hallucination rate). Hall. = expert-confirmed hallucination rate.

### 3.3 Trap Detection

Trap detection varied substantially across the 10 prespecified categories (Figure 2). Three categories achieved *>*98% overall detection: T02 (metformin at full dose with eGFR*<*30; 100%), T06 (PPI without indication; 98.6%), and T01 (NSAID in CKD; 98.3%). Intermediate detection was observed for T04 (gabapentin/pregabalin dose adjustment; 95%), T03 (DOAC dose adjustment; 95%), and T09 (tacrolimus–azole interaction; 92%, limited to 5 transplant cases). In contrast, two traps were poorly detected across all models: phosphate binder administration timing (T08; 10.9%, range 0–18%) and the hyperkalemia combination trap (T05; 32.5%, range 12.5–62.5%). Statin gap detection (T10) averaged 71.7% with the widest inter-model spread (33% for DeepSeek R1 to 100% for Grok 4.1 Fast). The poorly-detected traps (T08, T05) require integration of administration context or multi-drug risk assessment rather than single-drug contraindication lookup.

**Figure 1.**
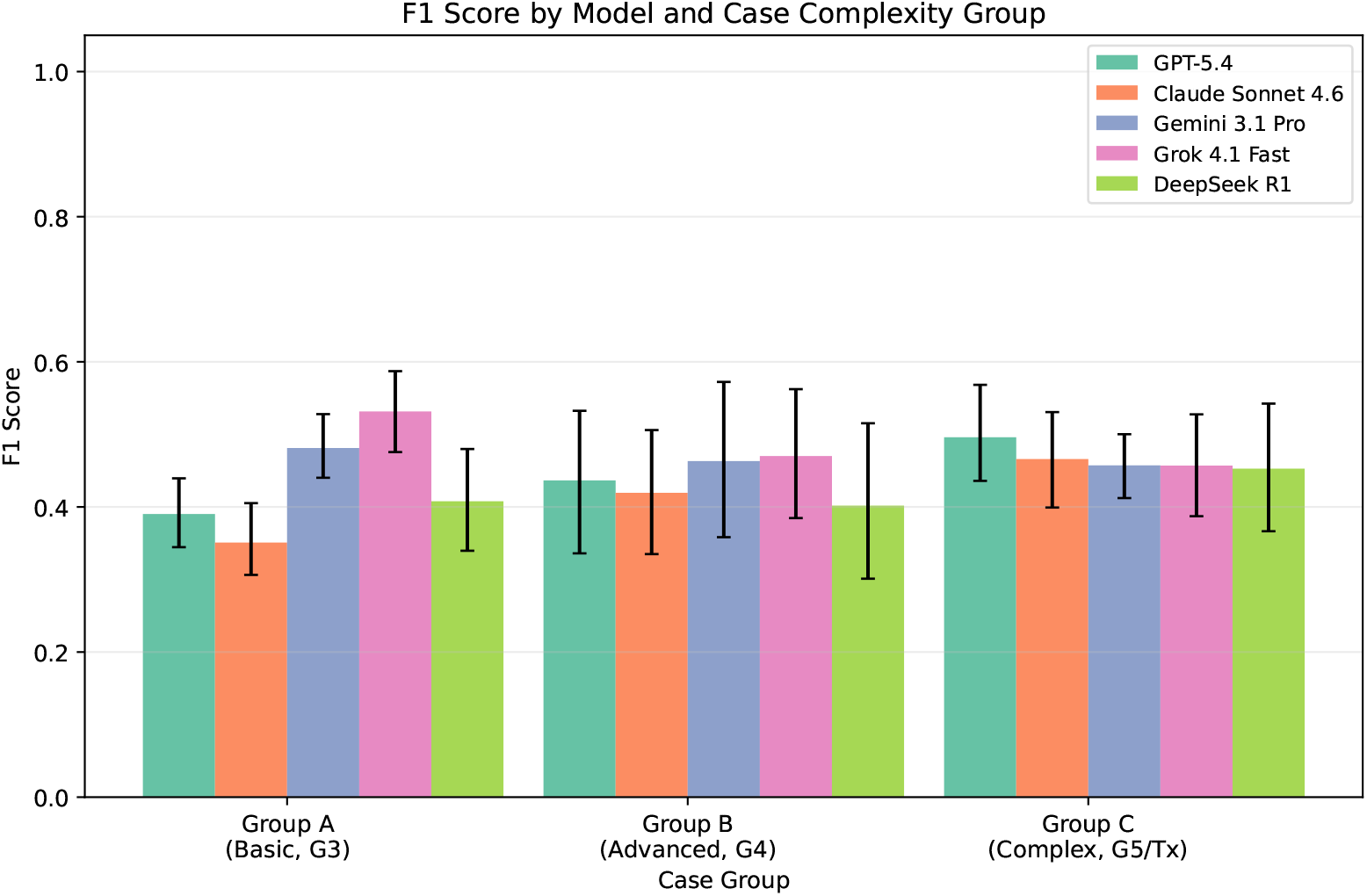
Macro F1 score by model and CKD complexity group. Bars represent per-case mean F1; error bars indicate 95% bootstrap confidence intervals (*n* = 2,000 resamples). Group A: CKD G3a–G3b (*n* = 20); Group B: G4 (*n* = 15); Group C: G5/G5D/transplant (*n* = 15). Friedman test was significant for Group A (*P <* 0.001) but not for Groups B (*P* = 0.114) or C (*P* = 0.334).

**Figure 2.**
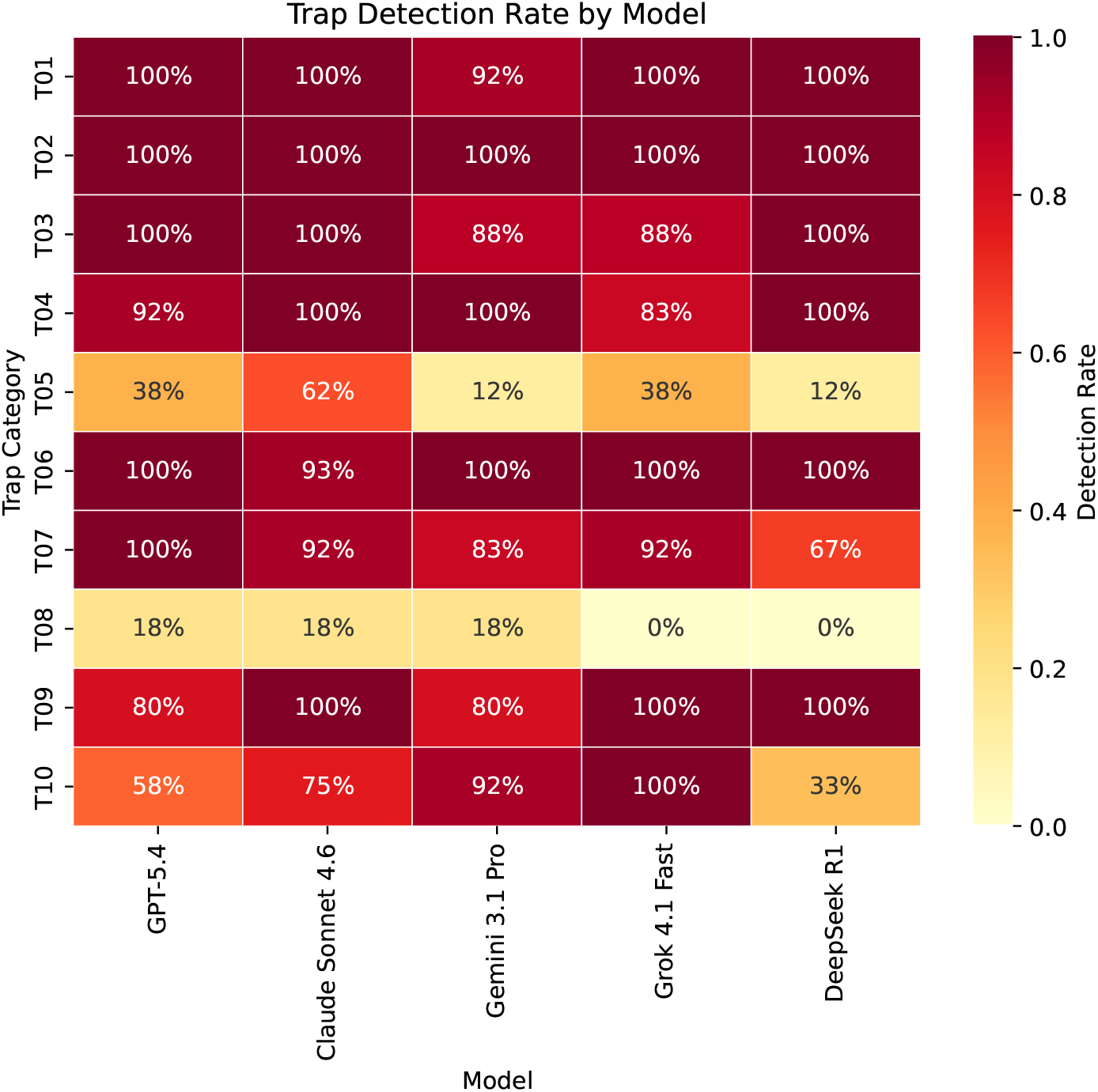
Trap detection rates by trap category and model. Rows represent 10 prespecified clinical trap types (T01–T10); columns represent five evaluated LLMs. Cell values indicate the proportion of cases in which the trap was correctly identified (consensus across 3 runs). Darker shading indicates higher detection. T08 (phosphate binder timing) and T05 (K^+^-sparing + ACEi/ARB combination) showed the lowest overall detection (≤33%).

### 3.4 Exploratory Upper-Bound Sensitivity Analysis (DR-06)

The PI classified all 1,759 false positives across five models by clinical validity on aggregated (drug, issue_type) patterns (*n* = 162 unique patterns): 1,637 (93.1%) were clinically valid recommendations beyond gold-standard scope (DR-06), 57 (3.2%) theoretically correct but clinically insignificant (DR-07), and 65 (3.7%) true clinical errors. Under this exploratory framework, extended *micro* F1 (TP/FP/FN aggregated across all 50 cases) increased to 0.66 (DeepSeek R1) – 0.82 (GPT-5.4; Table 3); the corresponding strict micro F1 ranged 0.41– 0.49 (note that strict *macro* F1 in Table 2 happens to span the same numerical range due to per-case averaging). An independent second nephrologist classified a 30% stratified sample; three-category agreement was 71% (*κ* = 0.21, fair), with 6 additional patterns reclassified as true errors, confirming that DR-06 boundaries are inherently subjective (Supplementary Figure S2). Extended F1 should be interpreted strictly as an exploratory upper-bound estimate; true performance likely falls between strict and extended micro F1. Demarcating clinically valid recommendations from gold-standard omissions in open-ended generation tasks represents a methodological boundary requiring prespecified ontologies in future benchmark iterations.

**Table 3.** Exploratory upper-bound sensitivity: strict versus extended F1.

|  | GPT-5.4 | Claude Sonnet 4.6 | Gemini 3.1 Pro | Grok 4.1 Fast | DeepSeek R1 |
| --- | --- | --- | --- | --- | --- |
| Strict F1 (micro) | 0.44 | 0.41 | 0.47 | 0.49 | 0.43 |
| Extended F1 (micro) | 0.82 | 0.78 | 0.73 | 0.71 | 0.66 |
| Total FP, $n$ | 459 | 525 | 273 | 245 | 257 |
| True FP, $n$ (%) | 10 (2.2) | 28 (5.3) | 3 (1.1) | 14 (5.7) | 10 (3.9) |
| DR-06, $n$ (%) | 441 (96.1) | 469 (89.3) | 257 (94.1) | 228 (93.1) | 242 (94.2) |
| DR-07, $n$ (%) | 8 (1.7) | 28 (5.3) | 13 (4.8) | 3 (1.2) | 5 (1.9) |

Total FP = sum across all model outputs prior to reclassification. True FP% = true errors / total FP per model. DR-06: clinically valid recommendations beyond gold-standard scope. DR-07: theoretically correct but clinically insignificant. Extended F1 excludes DR-06 and DR-07 from false positive count. **Exploratory analysis only:** DR-06/DR-07 classification was single-scorer post-hoc; independent second-reviewer validation on 30% of patterns showed only fair agreement ( $\kappa = 0.21$ ). Extended F1 is an upper-bound estimate and should not be interpreted as a performance estimate.

### 3.5 Hallucination

PI-adjudicated confirmed hallucination ranged from 0% (GPT-5.4) to 54% (DeepSeek R1; 27/50 cases), with Grok 4.1 Fast at 52% (26/50), Claude Sonnet 4.6 at 18%, and Gemini 3.1 Pro at 12%. A small number of cases remained pending adjudication at analysis lock (Claude *n* = 4, Grok *n* = 3, Gemini *n* = 1); conservative confirmed-plus-pending rates would be Claude Sonnet 4.6 22%, Grok 4.1 Fast 58%, Gemini 3.1 Pro 14% (DeepSeek R1 and GPT-5.4 unchanged). Only GPT-5.4 produced zero confirmed hallucinations across all 50 consensus evaluations. Hallucination patterns differed qualitatively: Grok 4.1 Fast’s were dominated by fabricated guideline section numbers and misattributed trial names, whereas DeepSeek R1 exhibited systematic pharmacological errors — a fabricated “losartan max 50 mg/day in CKD” dose cap (11 cases) and a non-existent “KDIGO 2023 bicarbonate guideline” citation (11 cases) — patterns clinically more dangerous as they could directly influence prescribing decisions [25].

### 3.6 Safety-Critical Errors

SCE rates (cases with ≥1 Level 3–4 missed problem) ranged from 22% (GPT-5.4 and Claude Sonnet 4.6; 95% CI: 13–35%) to 48% (DeepSeek R1; 95% CI: 35–61%), with significant heterogeneity (Cochran’s Q = 23.10, *P* = 0.0001; Figure 3). Significant pairwise differences (Bonferroni *α* = 0.005) were confirmed for GPT-5.4 vs DeepSeek R1 (*P* = 0.0009) and Claude Sonnet 4.6 vs DeepSeek R1 (*P* = 0.0009). Notably, GPT-5.4, Claude Sonnet 4.6, and Grok 4.1 Fast had low SCE rates (22%, 22%, and 28%) despite markedly different hallucination profiles (0%, 18%, 52% respectively), suggesting that safety-critical omissions and hallucinations were dissociated at the model level rather than tracking together (Supplementary Figure S1); a formal statistical test of independence was not prespecified.

**Figure 3.**
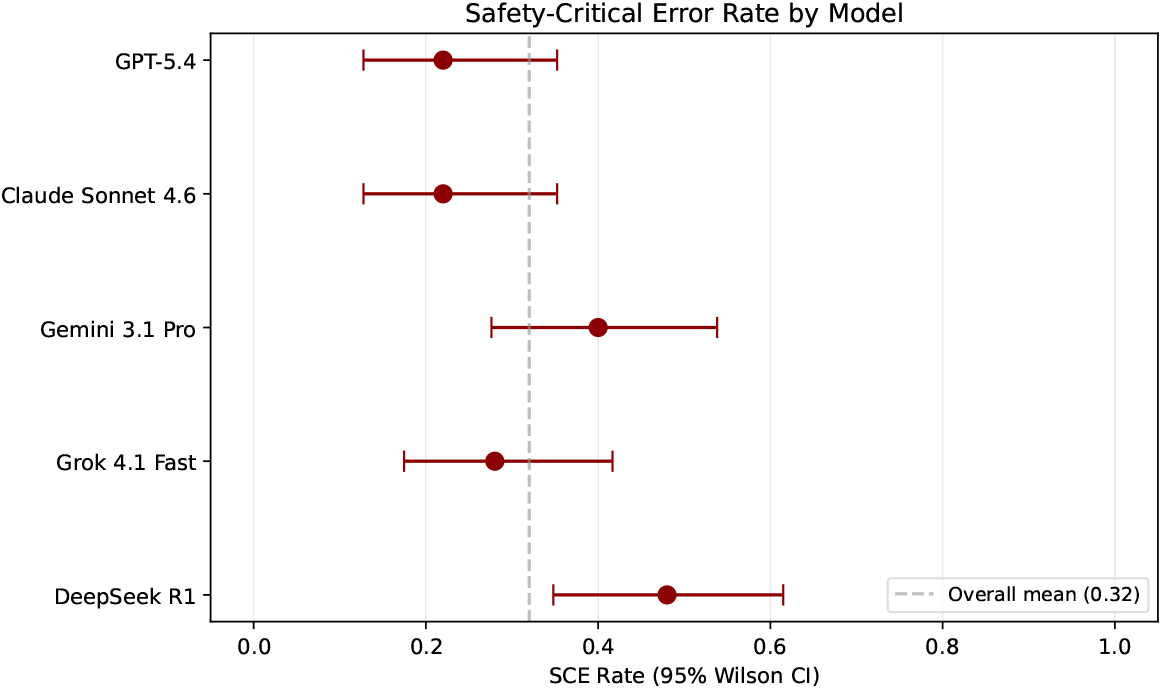
Safety-critical error (SCE) rates by model with 95% Wilson confidence intervals. SCE is defined as cases containing ≥1 missed problem classified as Level 3 (could cause harm) or Level 4 (contraindicated). Cochran’s Q test indicated significant heterogeneity across models (*Q* = 23.10, *P* = 0.0001). Vertical dashed line: overall mean SCE rate.

**Figure 4.**
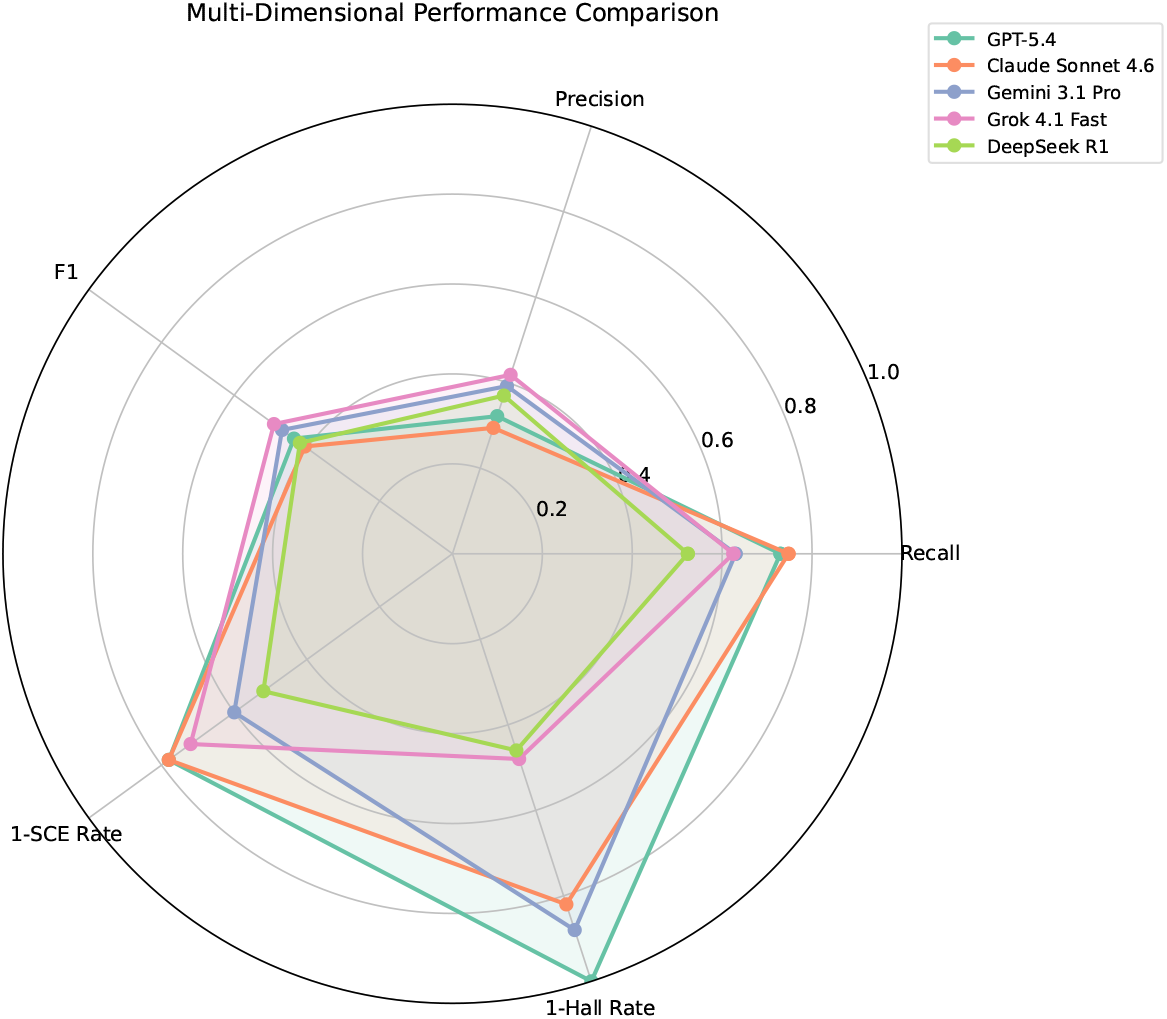
Multi-dimensional performance comparison across five models. Axes represent: Recall, Precision, F1, 1 − SCE rate (safety), and 1 − Hallucination rate (trustworthiness), each scaled 0–1. Larger enclosed area indicates better overall performance. GPT-5.4 shows the best safety–trustworthiness profile (zero hallucination, lowest SCE) despite moderate F1. Grok 4.1 Fast achieves highest F1 but is penalized by high hallucination (52%).

### 3.7 Intra-Model Consistency

Mean pairwise Jaccard similarity ranged from 0.48 ± 0.09 (DeepSeek R1 and Claude Sonnet 4.6) to 0.76 ± 0.15 (Gemini 3.1 Pro). Full three-run agreement was achieved in 0% of cases for four of five models; only Gemini 3.1 Pro achieved any full agreement (12%, 6/50 cases). These results demonstrate that under the tested API serving conditions, temperature-0 settings did not produce identical extracted clinical problem sets.

### 3.8 Subgroup Analysis

Within Group A (*n* = 20), Friedman test showed significant model differences (*χ*^2^ = 30.74, *P* < 0.001), with Grok 4.1 Fast performing best (F1 = 0.53; 95% CI: 0.48–0.59) and Claude Sonnet 4.6 lowest (F1 = 0.35; 95% CI: 0.31–0.41). Group C showed compressed inter-model separation (range 0.45–0.50); Groups B and C showed no significant inter-model differences (*P* = 0.11 and *P* = 0.33, respectively), though these analyses were underpowered with only 15 cases per group. Transplant subset (*n* = 5) results are in Supplementary Table S5.

### 3.9 Gold-Standard Reliability

Blinded inter-rater reliability between two nephrologists was *κ* = 0.934 (95% CI: 0.86–1.00; *n* = 92 problems across 15 cases; 96.7% agreement; Table 4) [26, 27].

**Table 4.** Inter-rater reliability: blinded scoring agreement.

| | Round 1<br>($n = 28$) | Round 2<br>($n = 50$) | All rounds<br>($n = 92$) |
| --- | --- | --- | --- |
| TP–TP (both agree detected) | 18 | 25 | 50 |
| FN–FN (both agree missed) | 8 | 24 | 39 |
| TP–FN (primary TP, second FN) | 2 | 1 | 3 |
| FN–TP (primary FN, second TP) | 0 | 0 | 0 |
| Agreement | 92.9% | 98.0% | 96.7% |
| Cohen’s $\kappa$ | 0.837 | 0.960 | 0.934 |
| 95% CI | 0.62–1.00 | not estimable<sup>a</sup> | 0.86–1.00 |

Blinded design: second nephrologist scored without access to primary scorer’s decisions to avoid anchoring bias.
Fifteen case-model pairs (6A+5B+4C) sampled across three rounds (6 pairs in Round 1, 7 in Round 2, 2 in Round 3), covering 30% of cases stratified by CKD group. Models balanced at 3 pairs each. Round 3 ( $n = 14$ ) achieved perfect agreement ( $\kappa = 1.0$ ) and is included in the “All rounds” column. <sup>a</sup>Round 2 standard error approached zero (only one disagreement among 50 items), causing the asymptotic 95% CI formula to fail; the all-rounds CI (0.86–1.00) is reported as the primary reliability estimate.

## 4 Discussion

A nephrology-specific trap-embedded benchmark was applied to evaluate five current-generation LLMs on open-ended comprehensive medication review—a task format that exposes failure modes poorly assessed by conventional multiple-choice formats. Three principal findings emerged. First, strict macro F1 ranged from 0.41 to 0.49, indicating that detection remained limited across all models despite their reported general-medicine capabilities. Second, in the micro-aggregated exploratory sensitivity analysis, F1 increased from 0.41–0.49 (strict) to 0.66–0.82 (extended) after excluding DR-06/DR-07 items, suggesting that many apparent false positives were not true errors, but rather plausible recommendations beyond the predefined gold-standard scope — a finding that requires independent confirmation given the subjective DR-06 boundaries (*κ* = 0.21 between reviewers; Supplementary Figure S2). True model performance likely lies between these bounds; the extended F1 reclassification is exploratory and hypothesis-generating only, and should not be interpreted as evidence of higher comparative performance. Third, safety profiles were markedly heterogeneous: hallucination ranged from 0% (GPT-5.4) to 54% (DeepSeek R1), and SCE rates from 22% to 48%, highlighting that model selection and guardrail design may affect downstream clinical risk.

The nephrology-specific blind spots identified here are unlikely to be captured by multiple-choice general-medicine benchmarks. Phosphate binder administration timing (T08; 11% detection) and the hyperkalemia combination trap (T05; 33%) require integration of administration context or multi-drug risk assessment rather than single-drug contraindication lookup. Similarly, statin gap detection (T10; 68%) reflects the nuanced KDIGO recommendation against statin *initiation* in dialysis patients, which several models failed to distinguish from statin omission. These findings argue for specialty-specific evaluation frameworks that probe domain knowledge beyond what multiple-choice formats can assess.

The highest strict F1 observed (0.49, Grok 4.1 Fast) is comparable to the top LLM performance reported by Yang et al. in general pharmacy prescription review [28], yet CKD-specific evaluation imposes harder tasks: nephrology-specific traps and open-ended format without predefined answer choices. Prior evaluation of ChatGPT on renal dose interventions yielded only 16.7% accuracy [19]; our GPT-5.4 recall of 0.73 suggests substantial generation-over-generation improvement, though direct comparison is confounded by task format differences.

A precision–recall tradeoff was apparent across models. GPT-5.4 (recall 0.73) and Claude Sonnet 4.6 (recall 0.75) showed higher recall but lower precision (0.29–0.32), consistent with longer responses flagging a larger set of candidate problems; Grok 4.1 Fast and Gemini 3.1 Pro showed lower recall (0.62–0.63) but higher precision (0.39–0.42), consistent with more selective reporting. The preferable operating point depends on the intended downstream use — prioritising sensitivity for screening, or precision for actionable alerts — and this study did not formally adjudicate between them. DeepSeek R1 hallucinations were qualitatively more dangerous than Grok 4.1 Fast’s fabricated citation numbers: systematic pharmacological errors (fabricated dose cap, non-existent guideline reference) in 11 cases each could plausibly influence prescribing without triggering clinical suspicion. This underscores the need for domain-specific guardrails — such as real-time cross-referencing against formulary databases or guideline APIs — before LLM-generated recommendations enter clinical workflow [29, 30].

Intra-model consistency (Jaccard 0.48–0.76) despite temperature-0 settings highlights an underappreciated challenge for clinical deployment. Non-determinism at the API level means identical prompts may yield different clinical recommendations across runs—even when deterministic output is explicitly requested. For Claude Sonnet 4.6, the lowest-consistency model (Jaccard 0.48), the same medication list could generate meaningfully different problem sets depending on query timing. For clinical decision support applications requiring reproducible outputs—a prerequisite for regulatory certification and clinical liability frameworks—this variability represents a design constraint that temperature settings alone cannot resolve.

Three limitations warrant consideration. First, all cases were synthetic; while constructed to reflect realistic CKD scenarios with guideline-referenced medications, real-world medication lists involve greater heterogeneity and contextual ambiguity. Specifically, real-world CKD polypharmacy frequently exceeds the 8–12 medications simulated here, and includes undocumented adherence variability, patient-specific dose modifications, and over-the-counter or herbal supplement use that were not represented. Synthetic-case performance should therefore be interpreted as a lower bound of the complexity that real-world cases impose. No direct human-performance comparator was included, precluding LLM-versus-clinician comparison; the absolute clinical meaning of the observed F1 and SCE values therefore cannot be benchmarked against nephrologist or pharmacist performance on the same cases. A prospective Phase B validation with real patient data and a human comparator arm is planned. Second, primary scoring was performed by a single nephrologist with model identity visible during Stage 2 adjudication, raising the possibility of unintentional scorer bias; blinded inter-rater reliability was *κ* = 0.934 (*n* = 92 problems), and the prespecified decision rules limit scorer subjectivity, but multi-center adjudication panels with full reviewer blinding would further strengthen construct validity. Third, DR-06 false-positive classification was conducted post-hoc; partial blinding via aggregated patterns mitigates but does not eliminate PI leniency bias, as confirmed by the second reviewer reclassifying 6 additional patterns as true errors. Extended F1 should be interpreted strictly as an upper bound.

This pilot synthetic benchmark, structured around ten categories of embedded clinical traps, surfaces failure patterns not typically captured by multiple-choice general-medicine assessments. The persistent miss of administration-context traps (9%) and multi-drug interaction patterns (28%) across all five tested models indicates that specialty-specific evaluation should precede clinical workflow use of LLM-assisted medication review. Heterogeneous hallucination (0–54%) and safety-critical error rates (22–48%) further underscore that model selection and domain-specific guardrail design are essential considerations for responsible deployment. Given the synthetic case basis, configuration asymmetries across models, and the exploratory nature of DR-06 reclassification, the inter-model comparisons reported here should be regarded as a hypothesis-generating comparative snapshot rather than a definitive leaderboard; prospective validation with real patient data and human comparators remains necessary before specific deployment recommendations can be made.

## Acknowledgments

The authors thank the nursing and pharmacy staff of the Division of Nephrology, An Nan Hospital, China Medical University, for clinical context during case construction.

## Data Availability Statement

All 50 synthetic cases, gold-standard answer files, 13 scoring decision rules, 4 hallucination rules, the standardized system prompt, all 750 raw LLM outputs, scored data files, and analysis code are available from the corresponding author on reasonable request. The study protocol (including decision rules) was version-controlled and finalized before scoring began (April 25, 2026). No real patient data were used in this study.

## Authors’ Contributions

Clinical case design validity review, manuscript drafting and revision: K-C.C.; research idea and study design: H-M.L.; case construction: H-M.L.; gold-standard development: H-M.L., H-J.L.; LLM testing and data acquisition: H-M.L.; scoring pipeline development and data analysis/interpretation: H-M.L.; statistical analysis: H-M.L.; inter-rater reliability assessment, gold-standard completeness review, and DR-06 independent adjudication (blinded second scorer): H-J.L.; critical manuscript review and proofreading: H-J.L., K-C.C. Each author contributed important intellectual content during manuscript drafting or revision and accepts accountability for the overall work by ensuring that questions pertaining to the accuracy or integrity of any portion of the work are appropriately investigated and resolved.

## Declaration of AI and AI-assisted technologies in the writing process

During the preparation of this work, the authors used Claude Code (Anthropic) and Codex (OpenAI) to assist with data analysis pipeline code development, proofreading, and translation. After using these tools, the authors reviewed, verified, and edited all content and take full responsibility for the content of the publication. All clinical case construction, gold-standard development, selection and inclusion of references, statistical analysis decisions, scoring adjudication, and scientific interpretation were performed and independently verified by the human authors.

