## Supplemental Tables for "Frontier Large Language Models for Comprehensive Medication Review in CKD Patients with Polypharmacy: A Trap-Embedded Synthetic Benchmark"

#### Contents


**Table S1. Trap Definitions** **3**

**Table S2. DR-06 Classification** **4**

**Table S3. Pairwise Tests** **5**

**Table S4. Sample-Size Context** **6**

**Table S5. Transplant Subset** **7**

**Table S6. Extraction Validation** **8**

**Table S7. Case Correction Sensitivity** **9**

**Figure S2. Strict vs Extended F1** **10**

**Appendix A. System Prompt** **11**

**Appendix B. Decision Rules** **12**

**Appendix C. Hallucination Rules** **13**

**Table S8. SCE Examples** **14**

**Figure S1. Error Distribution** **15**

**Table S9. Model Provenance** **16**

**Table S10. Individual-Run Performance Summary** **17**

**Appendix D. STROBE Checklist** **18**

### Table S1. Trap Category Definitions and Detection Rates

| ID | Description | Action | $n$ | GPT | Cld | Gem | Grk | DS | All |
|---|---|---|---|---|---|---|---|---|---|
| T01 | NSAID in CKD G3+ | Discontinue | 12 | 100 | 100 | 92 | 100 | 100 | 98 |
| T02 | Metformin full dose, eGFR$<$30 | Reduce/stop | 10 | 100 | 100 | 100 | 100 | 100 | 100 |
| T03 | DOAC standard dose, eGFR$<$30 | Adjust | 8 | 100 | 100 | 88 | 88 | 100 | 95 |
| T04 | Gabapentin not adjusted | Reduce dose | 12 | 92 | 100 | 100 | 83 | 100 | 95 |
| T05 | $K^+$-sparing + ACEi, eGFR$<$30 | Flag risk | 8 | 38 | 63 | 13 | 38 | 13 | 33 |
| T06 | PPI $>$8 wk, no indication | Deprescribe | 14 | 100 | 93 | 100 | 100 | 100 | 99 |
| T07 | Missing SGLT2i | Add | 12 | 100 | 92 | 83 | 92 | 67 | 87 |
| T08 | Phosphate binder timing | Flag admin | 11 | 18 | 18 | 18 | 0 | 0 | 11 |
| T09 | Tacrolimus + azole DDI | Flag DDI | 5 | 80 | 100 | 80 | 100 | 100 | 92 |
| T10 | Statin not prescribed | Add | 12 | 58 | 75 | 92 | 100 | 33 | 72 |

DS = DeepSeek R1. Values: detection rate (%). Each case contains $\geq$2 traps; 9 of 10 traps appear in $\geq$8 cases. T09 (tacrolimus–azole DDI) is limited to 5 transplant cases (C-11–C-15) by clinical design, as immunosuppressant interactions cannot be meaningfully embedded in non-transplant cases.

### Table S2. DR-06 False Positive Classification (Top 20 Patterns)

| Drug | Issue Type | $n$ | PI Classification |
|---|---|---|---|
| Aspirin | Deprescribing | 146 | DR-06 valid extra |
| Calcium carbonate | Deprescribing | 97 | DR-06 valid extra |
| Sodium bicarbonate | Missing therapy | 57 | DR-06 valid extra |
| Losartan | Monitoring needed | 55 | DR-06 valid extra |
| Furosemide | Dose adjustment | 53 | DR-06 valid extra |
| Furosemide | Deprescribing | 51 | DR-06 valid extra |
| Sodium bicarbonate | Dose adjustment | 47 | DR-06 valid extra |
| Ferrous sulfate | Dose adjustment | 43 | DR-06 valid extra |
| Hydrochlorothiazide | Deprescribing | 42 | DR-06 valid extra |
| ESA | Missing therapy | 40 | DR-06 valid extra |
| Lisinopril | Dose adjustment | 40 | DR-06 valid extra |
| Lisinopril | Monitoring needed | 40 | DR-06 valid extra |
| Calcitriol | Dose adjustment | 37 | DR-06 valid extra |
| Ferrous sulfate | Deprescribing | 37 | DR-06 valid extra |
| Furosemide | Monitoring needed | 34 | DR-06 valid extra |
| Ferrous sulfate | Monitoring needed | 32 | DR-06 valid extra |
| Ferrous sulfate | Drug interaction | 31 | DR-06 valid extra |
| Folic acid | Deprescribing | 30 | DR-06 valid extra |
| Vitamin D | Missing therapy | 29 | DR-06 valid extra |
| Spironolactone | Contraindication | 27 | DR-06 valid extra |

Showing top 20 of 162 unique (drug, issue_type) patterns. Total FP across all models: 1,759. DR-06 valid: 1,637 (93.1%). DR-07 theoretical: 57 (3.2%). True errors: 65 (3.7%). Classification was performed post-hoc by the PI. An independent second nephrologist classified a 30% stratified sample (49 patterns); three-category agreement was 71% ($\kappa = 0.21$), with the second reviewer classifying 6 additional patterns as true errors, consistent with mild PI leniency bias.

### Table S3. Pairwise Post-Hoc Comparisons

| Pair | Wilcoxon $W$ (F1) | $P$ | McNemar $\chi^2$ (SCE) | $P$ |
|---|---|---|---|---|
| GPT vs Claude | 416.0 | 0.051 | 0.07 | 0.789 |
| GPT vs Gemini | 392.0 | 0.044 | 4.92 | 0.027 |
| GPT vs Grok | 371.0 | 0.016 | 0.27 | 0.606 |
| GPT vs DeepSeek | 499.0 | 0.361 | 11.08 | <0.001* |
| Claude vs Gemini | 269.5 | <0.001* | 5.82 | 0.016 |
| Claude vs Grok | 275.5 | <0.001* | 0.57 | 0.450 |
| Claude vs DeepSeek | 523.0 | 0.505 | 11.08 | <0.001* |
| Gemini vs Grok | 441.0 | 0.277 | 2.50 | 0.114 |
| Gemini vs DeepSeek | 260.5 | 0.010 | 1.50 | 0.221 |
| Grok vs DeepSeek | 225.5 | <0.001* | 5.79 | 0.016 |

*Significant after Bonferroni correction ($\alpha_{\text{adj}} = 0.005$ for 10 pairs). Wilcoxon signed-rank for F1 (continuous); McNemar for SCE (binary).

### Table S4. Post-Hoc Sample-Size Context (Exploratory)

| Parameter | Value |
|---|---|
| Observed $\sigma$ (F1 SD, pooled) | 0.152 |
| Observed $\rho$ (mean inter-model correlation) | 0.624 |
| Observed $\Delta_{\max}$ (largest mean F1 difference) | 0.093 |
| $N$ (cases) | 50 |
| Minimum detectable $\Delta$ at 80% power | $\approx$0.055 |
| Required $N$ for $\Delta = 0.07$ (80% power) | 28 |
| Required $N$ for $\Delta = 0.05$ (80% power) | 55 |

Exploratory sample-size context using a two-sample paired approximation, not an exact Friedman power calculation. These estimates describe the detectable effect size given the observed variance and correlation structure. They should **not** be interpreted as confirmatory evidence strengthening the primary analysis result. $\sigma$: pooled within-model SD of per-case F1. $\rho$: mean pairwise inter-model correlation of per-case F1.

### Table S5. Transplant Subset (C-11 to C-15, $N = 5$)

| | GPT-5.4 | Claude | Gemini | Grok | DeepSeek |
|---|---|---|---|---|---|
| F1 (mean ± SD) | 0.50 ± 0.07 | 0.54 ± 0.10 | 0.47 ± 0.03 | 0.53 ± 0.18 | 0.62 ± 0.14 |
| Recall (mean) | 0.76 | 0.83 | 0.53 | 0.58 | 0.64 |
| Precision (mean) | 0.37 | 0.40 | 0.43 | 0.49 | 0.64 |
| Cases with SCE | 2/5 | 0/5 | 2/5 | 1/5 | 2/5 |
| Cases with halluc. | 0/5 | 2/5 | 0/5 | 1/5 | 3/5 |

$N = 5$; descriptive only, no confidence intervals computed. Transplant cases involve tacrolimus/MMF/prednisone with unique DDI and monitoring requirements (traps T09, T08). DeepSeek achieved highest F1 in this subset but also highest hallucination count; Claude Sonnet 4.6 (post-retest) achieved the highest recall (0.83) and zero SCE in this subset.

### Table S6. Extraction Validation by Model

| Model | Items Validated | Agreement | Notes |
|---|---|---|---|
| GPT-5.4 | 89 | 100% | — |
| Grok 4.1 Fast | 84 | 100% | — |
| DeepSeek R1 | 75 | 100% | — |
| Gemini 3.1 Pro | 67 | 100% | — |
| Claude Sonnet 4.6 | 18 | 100% | Extraction model is same family |
| **Total** | **333** | **100%** | Protocol target: $\geq$95% |

**Primary validation:** 30-case stratified random sample (12A + 9B + 9C). PI verified all (drug, issue_type, action) tuples against source LLM responses. Extraction model: Claude Sonnet 4.5 (Anthropic). No difference in accuracy between Claude-generated and other model responses. **Second validation:** An independent second nephrologist reviewed a separate stratified sample of 50 raw outputs (10 per model) using 500-character response previews plus extracted tuples; 49/50 agreed (98%). The single discrepancy involved a multi-drug issue where the pipeline captured one but not both drugs. This preview-based check may slightly overestimate extraction fidelity compared to full re-abstraction from raw outputs.

### Table S7. Sensitivity: Corrected-Case Subset Versus Full Cohort

| | Corrected cases ($n = 13$) | All cases ($n = 50$) |
|---|---|---|
| Cases affected | 13/50 (26%) | — |
| Correction type | SCr adjusted via CKD-EPI 2021 | — |
| Mean F1 (5-model average) | 0.433 | 0.443 |
| GPT-5.4 | 0.399 | 0.436 |
| Claude Sonnet 4.6 | 0.381 | 0.398 |
| Gemini 3.1 Pro | 0.486 | 0.469 |
| Grok 4.1 Fast | 0.497 | 0.491 |
| DeepSeek R1 | 0.400 | 0.419 |

13 cases had $>30\%$ SCr/eGFR mismatch corrected post-validation. eGFR, CKD stage, and gold standard remained unchanged; only SCr was adjusted to match the stated eGFR via CKD-EPI 2021. LLM testing was re-run on corrected cases. The corrected-case subset performed comparably to the overall cohort ($\Delta$F1 = 0.010), confirming stability of primary findings.

### Figure S2. Strict Versus Extended F1 Scores

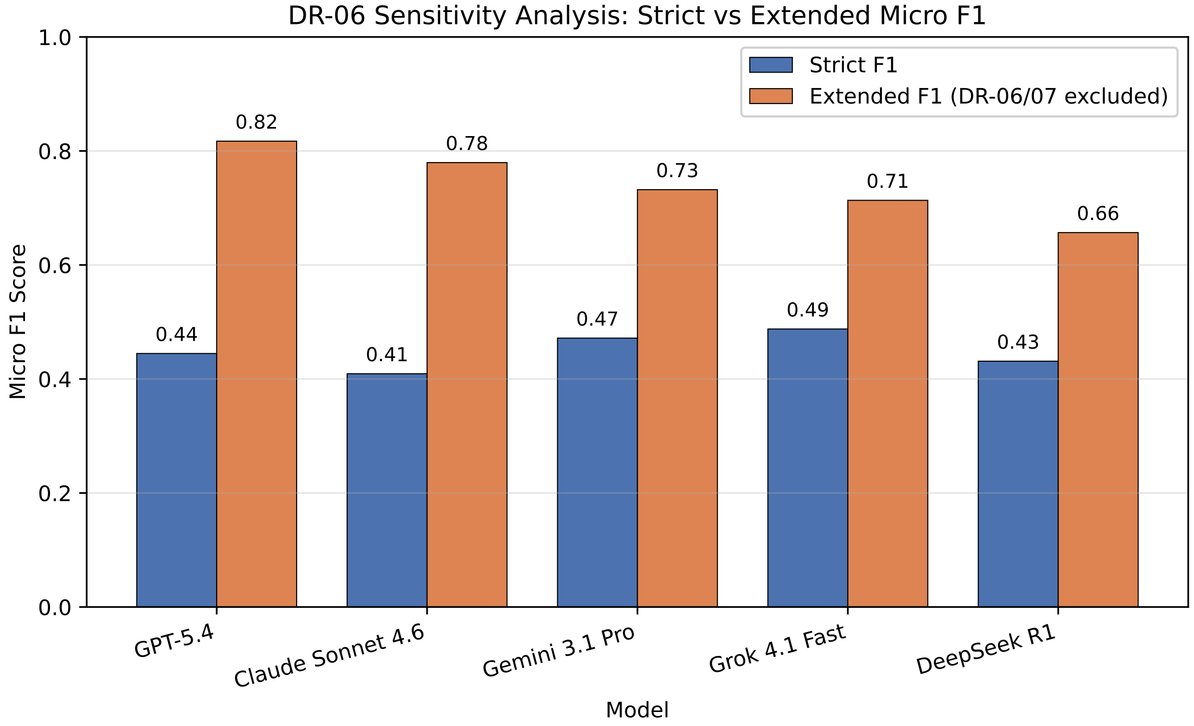


**Figure S2.** Strict versus extended (exploratory) F1 scores by model. Strict F1 (blue) counts all false positives equally. Extended F1 (orange) excludes clinically valid recommendations beyond gold-standard scope (DR-06) and theoretically correct but clinically insignificant items (DR-07). DR-06/DR-07 classification was performed post-hoc by the PI on aggregated patterns ($n = 162$). An independent second nephrologist classified a 30% stratified sample ($n = 49$); three-category agreement was 71% ($\kappa = 0.21$, fair), with the second reviewer classifying 6 additional patterns as true errors. Extended F1 therefore represents an upper-bound estimate; true performance likely falls between the strict and extended values. Delta values indicate the per-model F1 increase.

### Appendix A. Standardized System Prompt

The identical system prompt was used for all five models:

> *"You are a nephrology clinical pharmacologist conducting a comprehensive medication review for a CKD patient. Evaluate the entire medication list for: (1) dose adjustments needed based on renal function, (2) contraindicated medications, (3) drug-drug interactions, (4) deprescribing opportunities, (5) missing guideline-recommended therapies, and (6) administration/timing issues. For each issue identified, specify: the drug involved, the type of issue, and your recommended action with clinical reasoning."*

Full prompt text (including formatting instructions) available from the corresponding author on reasonable request.

#### Appendix B. Scoring Decision Rules (DR-01 through DR-13)

A True Positive requires ALL THREE criteria: (1) correct drug or pharmacological class, (2) correct issue type category, (3) correct clinical action direction.

| Rule | Summary |
|---|---|
| DR-01 | Correct drug, no specific dose $\rightarrow$ TP (detection primary) |
| DR-02 | Correct drug, wrong reasoning $\rightarrow$ TP |
| DR-03 | Correct drug, overly aggressive action $\rightarrow$ TP + additional FP |
| DR-04 | Two problems merged $\rightarrow$ 2 TPs if both addressed |
| DR-05 | One problem split $\rightarrow$ 1 TP (no double-count) |
| DR-06 | Valid problem not in gold standard $\rightarrow$ PI review (sensitivity) |
| DR-07 | Theoretical DDI without significance $\rightarrow$ FP Level 1 |
| DR-08 | Missing therapy, correct class wrong drug $\rightarrow$ TP |
| DR-09 | Partial DDI pair detection $\rightarrow$ FN |
| DR-10 | Deprescribing with wrong duration criterion $\rightarrow$ TP |
| DR-11 | Correct direction, dose outside range $\rightarrow$ TP |
| DR-12 | Monitor/consider only for actionable problem $\rightarrow$ FN |
| DR-13 | Statin in dialysis (KDIGO: don't initiate) $\rightarrow$ Not FN |

Full rule text with examples available from the corresponding author on reasonable request.

### Appendix C. Hallucination Rules (HR-01–HR-04)

| Rule | Scenario | Decision |
|---|---|---|
| HR-01 | Guideline reference with wrong/non-existent section | Hallucination |
| HR-02 | Correct guideline, paraphrased (no false specificity) | Not hallucination |
| HR-03 | Correct pharmacology, outdated recommendation | Not hallucination |
| HR-04 | Fabricated study with plausible details | Hallucination |

HR-03 note: incorrect pharmacology (e.g., fabricated dose caps) IS classified as hallucination under HR-03 when the mechanism/recommendation is factually wrong (not merely outdated). This distinction is critical for DeepSeek R1's systematic "losartan max 50 mg" pattern.

### Table S8. Representative Safety-Critical Errors (Level 3–4)

| Model | Level | Drug / Type | Missed Problem |
|---|---|---|---|
| GPT-5.4 | 4 | Spironolactone / DDI | $K^+$-sparing + ACEi at eGFR 17, $K^+$ 5.6 — hyperkalemia risk not flagged (B-01) |
| GPT-5.4 | 3 | Gabapentin / dose adj. | 900mg/day exceeds max at eGFR 52; should reduce to 300mg/day (A-07) |
| Claude | 4 | Spironolactone / DDI | $K^+$-sparing + ACEi at eGFR 18, $K^+$ 5.4 — not flagged (B-08) |
| Claude | 3 | Gabapentin / dose adj. | Not adjusted for CKD stage (B-02) |
| Gemini | 4 | Spironolactone / DDI | Same trap as above (B-01) |
| Gemini | 3 | Ibuprofen / contra. | NSAID in CKD G3a not discontinued despite triple whammy context (A-04) |
| Grok | 4 | Spironolactone / DDI | $K^+$-sparing + ACEi at eGFR 18 not flagged (B-08) |
| Grok | 3 | Gabapentin / dose adj. | 900mg/day at eGFR 45 not reduced (A-02) |
| DeepSeek | 4 | Spironolactone / DDI | Same trap (B-01) |
| DeepSeek | 3 | Ibuprofen / DDI | Triple whammy (NSAID+ARB+diuretic) not identified (A-01) |

Level 4: contraindicated combination with immediate harm potential. Level 3: clinically significant error requiring intervention. DDI = drug-drug interaction. The spironolactone + ACEi hyperkalemia trap (T05) was the most commonly missed Level 4 error across all models (overall detection: 33%).

### Figure S1. Error Level Distribution by Model

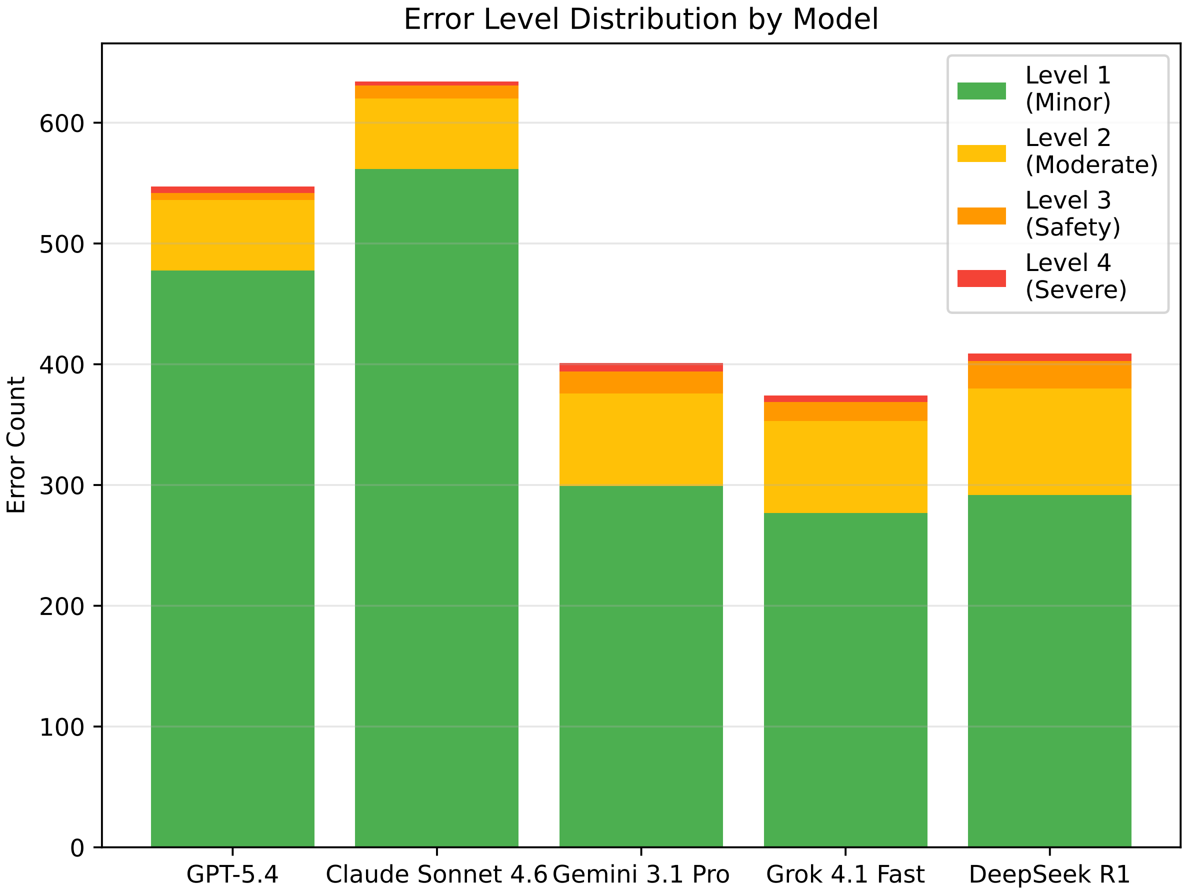


**Figure S1.** Distribution of false-negative error severity levels by model. Stacked bars show the proportion of missed problems at each severity level (Level 1: minor; Level 2: moderate; Level 3: safety-critical; Level 4: severe/contraindicated). Safety-critical errors (SCE; Levels 3–4) are highlighted. DeepSeek R1 had the highest proportion of Level 3–4 missed problems (48% of cases contained ≥1 SCE).

### Table S9. Model Provenance

| Study ID | API Model ID | Provider | Access | Run Dates | Reasoning Mode | Max Tokens |
|---|---|---|---|---|---|---|
| GPT-5.4 | gpt-5.4-2026-03-05 | OpenAI | Direct API | Apr 26–28 | Standard | 8,192 |
| Claude Sonnet 4.6 | anthropic/claude-sonnet-4.6 | Anthropic | OpenRouter[a] | May 17 | Standard | 16,384[a] |
| Gemini 3.1 Pro | gemini-3.1-pro-preview | Google AI | Direct API | Apr 26–28 | Standard | 8,192 |
| Grok 4.1 Fast | x-ai/grok-4.1-fast | xAI | OpenRouter | Apr 26–28 | Non-reasoning | 8,192 |
| DeepSeek R1 | deepseek/deepseek-r1 | DeepSeek | OpenRouter | May 3 | Reasoning | 8,192 |

All models: temperature=0, no tool use, no web search, and no study-specific safety filters or retrieval augmentation enabled. A single standardized system prompt (Appendix A) was used identically across all models. OpenRouter models accessed via the OpenRouter API; direct-API models accessed via provider-native endpoints. Grok 4.1 Fast was explicitly set to its non-reasoning configuration. DeepSeek R1 is an open-source model (January 2025 release) accessed through OpenRouter's hosted endpoint; exact checkpoint determined by OpenRouter routing at time of query. [a]Claude Sonnet 4.6 was originally tested April 26–28, 2026 via Anthropic Direct API at 8,192 max tokens, but exhibited output truncation at the token limit in 44/150 runs (29%; 60% in Group C). To eliminate this configuration confounder, Claude was re-tested on May 17, 2026 via OpenRouter (same underlying model) at 16,384 max tokens. All 150 retest outputs completed without truncation (max observed: 12,981 tokens; mean 7,877 tokens). All reported Claude results use the May 17 retest data. See Methods section "Provenance note: Claude retest at 16,384 tokens" for full rationale.

### Table S10. Individual-Run Performance Summary

| Model | Runs | F1 mean | F1 SD | F1 range | Recall mean | Precision mean |
|---|---|---|---|---|---|---|
| GPT-5.4 | 150 | 0.397 | 0.125 | 0.000–0.706 | 0.720 | 0.281 |
| Claude Sonnet 4.6 | 150 | 0.347 | 0.116 | 0.095–0.667 | 0.708 | 0.238 |
| Gemini 3.1 Pro | 150 | 0.444 | 0.154 | 0.125–0.875 | 0.604 | 0.363 |
| Grok 4.1 Fast | 150 | 0.455 | 0.151 | 0.143–0.824 | 0.616 | 0.373 |
| DeepSeek R1 | 150 | 0.391 | 0.166 | 0.000–0.889 | 0.535 | 0.318 |

Individual-run metrics summarize all 750 model outputs before consensus aggregation. Consensus-level results remain the primary analysis because the study prespecified three independent runs per case followed by majority voting.

### Appendix D. STROBE Checklist for Cross-Sectional Studies

| # | Item | Done? | Location |
|---|---|---|---|
| **Title and abstract** | | | |
| 1a | Study design in title/abstract | Yes | Abstract: “Cross-sectional evaluation” |
| 1b | Informative abstract | Yes | Structured abstract (KM 9-heading format) |
| **Introduction** | | | |
| 2 | Scientific background/rationale | Yes | Introduction ¶1–3 |
| 3 | Objectives | Yes | Introduction ¶4 |
| **Methods** | | | |
| 4 | Study design | Yes | Methods §Study Design |
| 5 | Setting | Yes | Methods §Synthetic Case Construction |
| 6 | Participants | Yes | Methods §Synthetic Case Construction (50 cases, 3 groups) |
| 7 | Variables | Yes | Methods §Gold Standard Development (problem types, severity) |
| 8 | Data sources | Yes | Methods §LLMs + §Scoring Pipeline |
| 9 | Bias | Yes | Methods §Scoring Pipeline (extraction bias check); Discussion Limitations |
| 10 | Study size | Yes | Methods (50 cases × 5 models × 3 runs = 750) |
| 11 | Quantitative variables | Yes | Methods §Statistical Analysis |
| 12 | Statistical methods | Yes | Methods §Statistical Analysis (Friedman, Cochran’s Q, bootstrap) |
| **Results** | | | |
| 13 | Participants | Yes | Results §Case Characteristics (Table 1) |
| 14 | Descriptive data | Yes | Table 1 (demographics, eGFR, medications) |
| 15 | Outcome data | Yes | Table 2 (F1, recall, precision, SCE, hallucination) |
| 16 | Main results | Yes | Results §Overall Model Performance |
| 17 | Other analyses | Yes | Results §DR-06 Sensitivity (exploratory), §Subgroup |
| **Discussion** | | | |
| 18 | Key results | Yes | Discussion ¶1 |
| 19 | Limitations | Yes | Discussion ¶5 (5 limitations) |
| 20 | Interpretation | Yes | Discussion ¶2–4 |
| 21 | Generalizability | Yes | Discussion Limitation #1 (synthetic cases) |
| **Other** | | | |
| 22 | Funding | Yes | Funding section |
